# Preferences and barriers to mental help seeking among students attending the University of Dschang, West region, Cameroon

**DOI:** 10.1101/2020.11.29.20214544

**Authors:** Martial Sonkoue Pianta, Linda Evans Eba Ze, Baame Esong Miranda, Rene Mih Tah, Benjamin Momo Kadia

## Abstract

**Background:** Mental disorders are a major source of disability and receive little attention in developing countries in general and particularly in Cameroon. Students are at a higher risk of developing mental disorders. The aim of this study was to assess students’ mental help seeking preferences and the barriers to mental health seeking.

**Methods:** We conducted a cross sectional study. Participants were selected using a multistage sampling method. A self-administered questionnaire was used to collect data on sociodemographic characteristics preferences and barriers to mental health seeking.

**Results:** Of the 84.23%[534/634, CI :81.18%- 86.86%] who consider depression a serious health problem 34.14%[183/534, CI : 30,25%- 38.26%] will go to a psychologist, in case of a mental health problem.. Most of the participants 47.32% [300/634,CI : 43,46%- 51.21%] prefer a private hospital for healthcare. The greatest barrier to health access is the inability to pay 38.64%[245/634, CI : 34.93%- 42.49%].

**Conclusion:** Interventions to increase awareness on available health services, implementation of universal health coverage and the promotion of mental health could greatly improve health seeking behaviour and access to mental healthcare.

## INTRODUCTION

Health-seeking behaviour has been defined as the activity undertaken by individuals who perceive themselves to have a health problem or to be ill for the purpose of finding an appropriate remedy[1].

Help-seeking behaviour has been conceptualized as a multi-stage process amongst mental-health researchers[2]. According to Eisenberg[2], individuals begin by experiencing a specific health problem, followed by recognizing a need for professional help. They then evaluate the costs and benefits of receiving treatment (within the context of social norms regarding seeking help), and take action to receive care by choosing one of several types of help for mental health concerns. Most mental health disorders emerge from age 15 to age 24 [3] and university students are at the age of early onset of mental illness [4]. Post-secondary students are at a fragile stage in their lives. The transition from high school to post-secondary education is characterized by change, adjustment, and ambiguity relating to a disruption of routines, security, predictability, and a loss of sense of control that was established during high school [5]. Post-secondary institutions aim to support students through various resources pertaining to mental health, academia, physical activity, and finances[2]. Despite the resources that exist on campuses, students’ mental health needs are still unmet[2]. While there is evidence that students have low help seeking rates[2,6], the majority of these students seek help from informal sources [7].

Mental disorders contributes largely to the burden of diseases worldwide, the prevalence of these disorders is higher among students than in the general population. Previous studies have shown a prevalence of major depressive disorder among Cameroonian nursing and medical students of 26.40% [8] and 30.6% [9] respectively. Furthermore primary care physicians have insufficient knowledge attitudes and practices about management of depression [10]. Without appropriate and timely treatment these disorders can lead to lifelong impairments and even suicide. Early recognition and provision of care can curb down the burden. There is insufficient provision of care and underutilisation of available services worldwide and more particularly in Cameroon. The objective of this study was to determine student’s preferences and barriers to seeking help for mental disorders.

## METHOD

### Ethical considerations

Ethical registry was granted by the Cameroon National Committee for Research Ethics for Human Health. The registry number is 2018/07/1081/CE/CNERSH/SP.

### Study design and period

This was a descriptive cross-sectional study conducted from the 10 March to the 30 April 2018.

### Study area and setting

The study was conducted on the main campus of the University of Dschang. The university of Dschang is one of the eight public universities of Cameroon. It is a bilingual university located in the West region of Cameroon in Menoua division within the town of Dschang and had a population of about 32000 students in 2018. The University of Dschang has six Faculties located on the main campus (Faculty of Science, Faculty of letters and human sciences, Faculty of economics and management, Faculty of law and political sciences, Faculty of agronomy and agricultural sciences, faculty of medicine and pharmaceutical sciences) and two institutions located out of the main campus (Foumban Institute of fine arts and Fotso Victor institute of Bandjoun). The faculty of medicine and pharmaceutical sciences was newly created by a presidential decree and went functional during academic year 2017 /2018. A small guidance counseling unit is located at the rectorate. Even though it serves the whole university, the unit is underutilized and seldom functional.

### Study participants and selection criteria

Students from the University of Dschang during the 2017 and 2018 academic year. The following selection were set:

#### Eligibility criteria

Being a registered student of the University of Dschang in the academic year 2017/2018.

#### Inclusion criteria

Being a registered student of the University of Dschang in the academic year 2017/2018 and consent to participate in the study.

#### Exclusion criteria

Refusal to participate; not responding to up to 80% of questions on the questionnaire.

### Sampling procedure

A multistage random sampling method was used. Two faculties were selected at random from the six faculties of the University of Dschang (FESM: Faculty of Economic Sciences and Management, FAAS: Faculty of Agronomy and Agricultural Sciences, FS: Faculty of Science, FLPS: Faculty of Law and Political Sciences FLHS: Faculty of Letters and Human Sciences, FMPS: Faculty of medicine and pharmaceutical sciences) and participants were recruited from strata designated by cycle of study in each selected faculty. The strata were the bachelors, masters and PhD study cycles. A systematic review found a prevalence of depression of 33% among students[11]. Using the Cochran’s sample size determination formula:

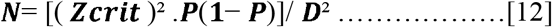

and substituting p=33%, relative precision (D) =0.05, normal standard deviate Zcrit=1.96 at significance criterion of 0.05, we obtained a minimum sample of N=339 students. Adjusting for a non-response rate of 10% and a design effect of 2 we got a final sample size of 758 students.

### Variables and data collection

A semi-structured questionnaire was conceived by the principal investigator and pre-tested on 50 students from the Faculty of Medicine and Biomedical Sciences and the Faculty of Science of the University of Yaounde 1. Participants were students from the Faculty of Agronomy and Agricultural Sciences (FAAS) and the Faculty of Economic Sciences and Management (FESM). After explaining the purpose of the study, participants’ consent was obtained before they responded to the self-administered questionnaire consisting of sociodemographic characteristics, preference for a healthcare provider, preference for an institution and barriers to healthcare. After collecting sociodemodemographic data, only participants who declared depression to be an important health problem were administered the rest of the questionnaire.

### Statistical analysis

Data were analysed using Epi-info version 7.2.2.6 statistical software. Descriptive statistics

(mean, frequencies and percentages) were used to report participants demographics, preference for a healthcare provider, preference for an institution and barriers to healthcare at a 95% confidence interval.

## RESULTS

### Sociodemographics

A total of 634 participants were included in the study. which 307 were females and 327 were males giving a total response rate of 84%. Most of the participants are single (97.2%) live single in a room (51%), are catholics (58.2%). 9% had a chronic disease, 0.04 were abused sexually, and 9% had a suicidal thought within the last 2 years. The majority of the participants parents had completed secondary education (45%).

**Table 1:** Sociodemographic characteristics

| Variable | Total | Female (307) | Male( 327) |
| --- | --- | --- | --- |
| **Lodging** |  |  |  |
| **Cohabitation in a room** | 75 (12%) | 40 (13%) | 35 (10.7%) |
| **Single in a room** | 324 (51%) | 200 (61.2%) | 92(28.1%) |
| **Family house** | 235 (37%) | 92 (28.13%) | 200(61.2%) |
| **Matrimonial status** |  |  |  |
| **Married** | 10 (1.6%) | 8 2.(6%) | 2 (0.6%) |
| **Divorced** | 3 (0.5%) | 3 (1%) | 0 |
| **Single** | 616 (97.2%) | 292 (95.1%) | 324 (99%) |
| **Widow(er)** | 5 (08%) | 4 (3%) | 1 (0.3%) |
| **Religion** |  |  |  |
| **Animist** | 2 (0.3%) | 1 (0.3%) | 1 (0.3%) |
| **Pentecostal** | 38 (6%) | 17 (5.5) | 21 (6.4%) |
| **Adventist** | 10 (1.6%) | 2 (0.7%) | 8 (2.5%) |
| **Catholic** | 369 (58.2%) | 196 (63.8%) | 196 (63.8%) |
| **Islamist** | 39 (6.15%) | 18 (5.7%) | 21 (6.4%) |
| **Jehovah Witness** | 7 (1.1%) | 5 (1.6%) | 2 (0.6%) |
| **Protestant** | 145 (22.9%) | 62 (20.2%) | 83 (25.6%) |
| **Other** | 24 (3.8%) | 6 (2%) | 18 (5.5%) |
| **Parents education** |  |  |  |
| **Primary** | 75 (11.8%) | 33 (10.8%) | 42 (12.8%) |
| **Secondary** | 285 (45%) | 144 (46.9%) | 141 (43.1%) |
| **Higher** | 274 (43.2%) | 130 (42.4%) | 144 (44%) |
| **Chronic disease** | 54 (9%) | 17 (6%) | 37 (11%) |
| **Sexual abuse** | 25 (0.04%) | 20 (6.5%) | 5 (1.5%) |
| **Suicidal thoughts within the last 2 years** | 57 (9%) | 31 (10.1%) | **26 (8%)** |

A total of 84.23% of the participants consider depression a serious health problem.

### Preference of healthcare provider and health institution in search of healthcare for a mental health problem

Participants will mainly prefer to seek help from a psychologist (34.08%), a medical doctor (32.21%) and others ie. Family and friends (20.79%) in case of a mental health problem. In terms of health institution, majority prefer a private hospital (50.19%) followed by the district hospital (39.7%) and only 5.06% will prefer the medical social center of the University in case of a mental health problem.

**Table 2:** Preferences and barriers to mental health seeking

| **Provider of help /care** | **Frequency(N=534)** | **Proportion(95% CI)** |
| --- | --- | --- |
| **Medical doctor** | 172 | 32.21(28 .39-36 .29) |
| **Pharmacy shop** | 15 | 2.81(1.71-4.58) |
| **Medicine vendor** | 5 | 0.94(0.40-2.17) |
| **Psychologist** | 182 | 34.08(30 .19-38 .20) |
| **Spiritual care** | 42 | 7.87(5.87-10 .46) |
| **Traditional healer** | 7 | 1.31(0.64-2.68) |
| **Other(Family, Friends)** | 111 | 20.79(17 .56-24 .43) |
| **Total** | 534 | 100 |
| **Institution** | **Frequency (N=534)** | **Proportion(95% CI)** |
| **District hospital** | 212 | 39.70(35.64-43.91) |
| **Medical social center** | 27 | 5.06(3.50-7.26) |
| **Private hospital** | 268 | 50.19(45.96-54 .41) |
| **Other** | 27 | 5.06(3.50-7.26) |
| **Total** | 534 | 100 |

### Barriers to health seeking

The main barriers to health seeking were the inability to pay (40%), ignorance of health services(19%), Knowing how to deal with depression(13%) and staff attitude(12%).

**Figure 1:**
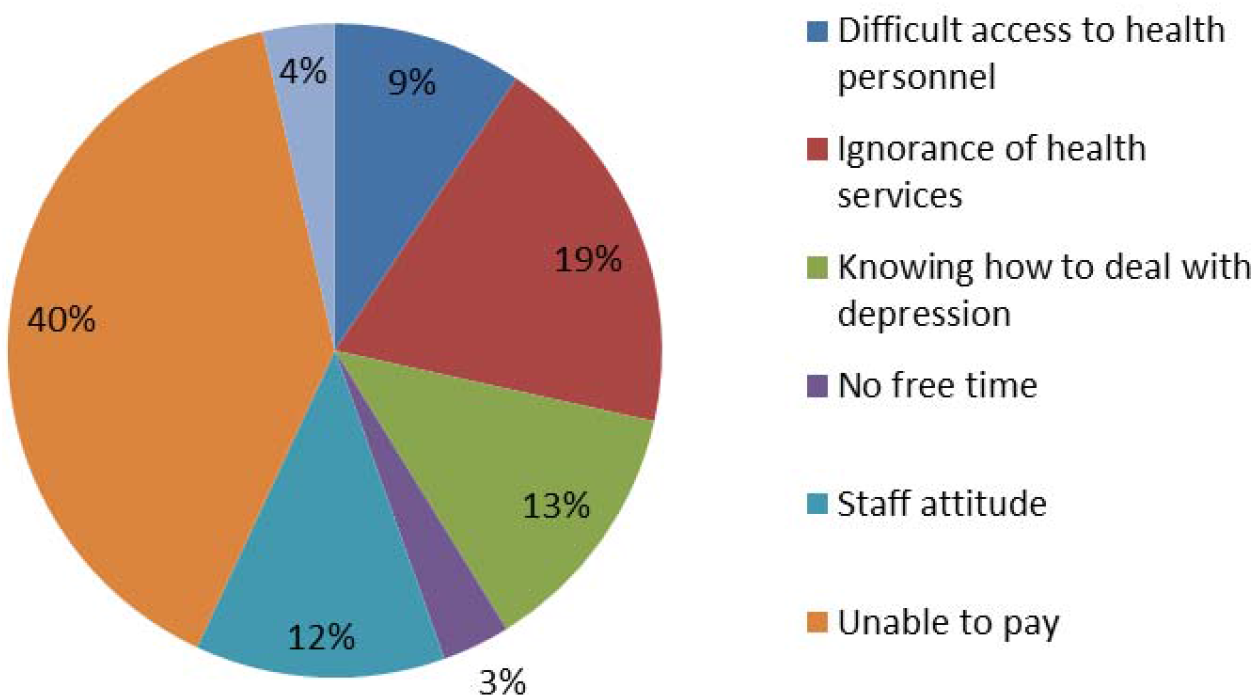
Frequency of barriers to healthcare access

## DISCUSSION

This study examined the preferences and barriers to seeking mental health among a sample of university students of a public university in Cameroon. The majority of participants consider depression a serious health problem. Psychologists, Medical doctors and others (Family and friends) are the main providers of help/ care participants will prefer to seek help/care from in case of a mental health problem. Concerning the institution, Participants prefer a private hospital. The main barriers to seeking help are the inability to pay, ignorance of health services and knowing how to deal with depression. The strengths of this study include the demographic profile which is representative of the national university student population, the multistage random sampling and the large sample size.

It was found that the most preferred healthcare/help provider in case of mental health problem include Psychologists (34.08%), medical doctors(32.21%) and others(family and friends) (20.79%). While this shows a predominant preference for trained mental health professional this contrasts with results from two previous studies in Ethiopia, one among students at Jimma university and the other among urban residents in Southeast Ethiopia where the majority of participants (83.8% of students) and (83% of urban residents) respectively who sought help for a mental health problem did so from informal sources [7,13]. Contrasting results were equally obtained in another study in Nigeria [14] with only 1.5% of partcipants(teachers) recommending help from professional sources (psychiatrists or psychologists). It should be noted that in our study, participants reported their preferences which may differ from their actual help seeking practices and could explain the contrasting results from other studies where participants reported their past sources of help. Differences in level of health literacy could as well explain discrepancies observed with other studies[15]. The emerging pathways to psychiatric care underline the importance of improving awareness through campaigns that will facilitate the recognition of psychiatric disorders [16].

Only 5.06% of participants preferred the medical social center whose role is to provide healthcare to students. This concurs with a study by Afolabi [17] who found that students sought help from community pharmacies and their peers from related academic disciplines than from the university health center. While 39.70% of the participants would prefer the District hospital which is the main public hospital in the area a greater proportion of participants will prefer a private hospital (50.19%). These findings align with another study in Oman[18]. In private hospitals patients could easily approach anyone including the reception staff and all are helpful, and the private hospitals are equipped with modern equipment, and doctors treat patients in a friendly manner[18].

Inability to pay (39.51%) and ignorance of health services (19.10%) were the main barriers to seeking healthcare. Similar results were obatained in Nigeria[17]. Most of the specialist centres where highly trained personnel work are in urban areas and for a large part of the population access to them is limited by distance and cost[19]. The implementation of universal health coverage and the promotion of available mental health services could significantly improve access to mental healthcare. More efforts should equally be made towards strengthening the health system as a whole.

This study has limitations. First the assessment of preference which may differ from the actual health-seeking behavior. Secondly stigma which is a barrier to mental help seeking was not assessed so results from this study should be interpreted with caution.

## CONCLUSION

Participants prefer formal sources when seeking mental health. Most of the participants prefer a private hospital when seeking for mental health. There is a little preference for the medical social center. The main barriers to seeking mental health were the inability to pay, ignorance of health services and staff attitude. Further research should assess actual mental health seeking behavior and the effect of stigma on mental health access of students at a larger scale.

## Data Availability

The data that support the findings of this study are avalaible from corresponding author upon reasonable request

## Author Contributions

SPM conceived and designed the study. SPM, LEEZ, BEM and MRT collected the study data. SPM, BMK analysed and interpreted the data. SPM, BMK, LEEZ, BEM and MRT prepared the original manuscript. All authors read and approved the final manuscript

